# Modular Robotic NOSES-II for Mid-Rectal Cancer: A Preliminary Feasibility Study of a Structured Surgical Procedure

**DOI:** 10.64898/2026.08.15.26359890

**Authors:** Ping Liu, Linsen Zhang, Kun Yu, Xianglong Lu, Wenliang Li

**Author notes:** Corresponding author: Wenliang Li, M.D., Ph.D. Department of Gastrointestinal Surgery, The Second Affiliated Hospital of Kunming Medical University, No. 374 Dianmian Avenue, Wuhua District, Kunming, Yunnan Province 650033, China.

## Abstract

**Background and Objectives:** Robotic-assisted natural orifice specimen extraction surgery (NOSES) is a minimally invasive approach for mid-rectal cancer, but it is technically complex and lacks a standardized operational framework. This study aimed to propose and preliminarily validate a structured modular robotic NOSES-II surgical procedure for mid-rectal cancer.

**Methods:** This was a retrospective observational study that consecutively enrolled 11 patients with mid-rectal cancer who underwent modular robotic NOSES-II surgery at our center between December 2024 and August 2025. All procedures were performed by the same surgical team strictly following the predefined six-module structured surgical protocol. Perioperative indicators, pathological outcomes, and patient-reported outcomes (PROs) at 4–8 months postoperatively were collected. Key techniques were analyzed via high-definition surgical videos.

**Results:** All 11 procedures were completed successfully without conversion to open surgery. The mean operative time was 299.6 ± 54.2 minutes, and the mean intraoperative blood loss was 94.1 ± 50.6 mL. The mean number of harvested lymph nodes was 17.4 ± 8.4, with a 100% R0 resection rate. No severe complications (Clavien-Dindo grade ≥ III) occurred. The mean postoperative hospital stay was 8.2 ± 1.5 days. Postoperative PROs indicated good defecatory function, urinary function, and overall quality of life.

**Conclusions:** Preliminary findings demonstrate that the structured modular robotic NOSES-II procedure is safe and feasible for the treatment of mid-rectal cancer. This modular protocol provides a clear and reproducible technical framework for this complex procedure, and it is expected to achieve favorable functional preservation while ensuring oncological radicality.

## 1. Introduction

Colorectal cancer remains one of the major global health burdens[1–3]. For surgery of mid-low rectal cancer, surgeons must strike a balance between radical resection and organ function preservation due to the complex anatomical environment[4–6]. Total mesorectal excision (TME) is the current standard procedure[7–9]. The combination of robotic surgical systems and natural orifice specimen extraction surgery (NOSES) provides new possibilities for more precise and minimally invasive radical resection of rectal cancer[10,11]. The three-dimensional high-definition vision and flexible instruments of the robotic system facilitate fine dissection, neurovascular protection, and reduction of postoperative complications in the narrow pelvis[12–14]. Meanwhile, NOSES technology further achieves minimal invasiveness by avoiding abdominal auxiliary incisions, improving cosmetic outcomes while reducing postoperative complications[15,16].However, robotic NOSES-II surgery involves complicated steps, has a high technical threshold, and lacks unified operational specifications, resulting in a steep learning curve that limits its widespread clinical application[17–19]. Decomposing complex surgeries into standardized modules is an effective strategy to improve their reproducibility, safety, and teachability[20–22]. At present, there is still no widely recognized structured scheme for robotic NOSES-II for mid-rectal cancer. Therefore, this study aims to systematically elaborate a structured modular robotic NOSES-II surgical procedure applicable to mid-rectal cancer for the first time, analyze its six core technical modules in detail via high-definition surgical videos, and preliminarily evaluate the perioperative safety, oncological outcomes, and patient-reported functional and quality-of-life outcomes of this protocol.

## 2. Methods

### 2.1 Study Design and Ethics

This was a single-center retrospective observational study. The protocol was approved by the Ethics Committee of the Third Affiliated Hospital of Kunming Medical University, Yunnan Cancer Hospital, Yunnan Hospital of Peking University Cancer Hospital (Approval No.: KYLX2026-250), and informed consent was obtained from all patients. The study was conducted in accordance with the principles of the Declaration of Helsinki, and reporting followed the STROBE guidelines.

### 2.2 Patient Population

Patients with mid-rectal cancer who underwent modular robotic NOSES-II surgery between December 2024 and August 2025 were consecutively enrolled.

- **Inclusion criteria:** (1) Pathologically confirmed rectal adenocarcinoma; (2) Tumor inferior margin 5–10 cm from the anal verge; (3) Clinical stage cT1–3N0–2M0; (4) Maximum tumor diameter < 3 cm; (5) Body mass index (BMI) < 30 kg/m².
- **Exclusion criteria:** (1) Distant metastasis detected on preoperative imaging or during surgery; (2) Requirement for emergency surgery; (3) Severe organ dysfunction; (4) History of pelvic radiotherapy.

### 2.3 Structured Surgical Procedure

The surgery was performed based on the principles of *“plane priority, sharp dissection, and nerve guidance”*, following six predefined modules: **Module 1:Medial approach and plane establishment:** The peritoneum was incised below the projection of the inferior mesenteric artery (IMA) to enter and expand the Toldt space and retrorectal space. The anterior fascia of the hypogastric nerve was used as the dorsal landmark to construct the “holy plane”.**Module 2:Management of the IMA region:** D3 lymph node dissection was performed at the root of the IMA, and the IMA was transected at a low position distal to the origin of the left colic artery (LCA) to preserve LCA blood supply.**Module 3:Mobilization of the left colon:** Combined medial Toldt space and lateral approach was used to fully mobilize the left colon and splenic flexure, with protection of the ureter and gonadal vessels.**Module 4:Mesenteric tailoring and blood supply assessment:** The mesentery was skeletonized and tailored along the descending branch of the left colic artery. Intraoperative indocyanine green (ICG) fluorescence imaging was used to assist in assessing intestinal blood supply.**Module 5:Circumferential rectal dissection:** Following TME principles, dissection was performed along the anatomical plane in the order of “posterior → lateral → anterior”, with preservation of the pelvic autonomic nerves throughout the procedure. **Module 6:Specimen extraction and reconstruction:** The NOSES-II B method was adopted. The specimen was extracted transanally, and intracorporeal end-to-end anastomosis was completed with seromuscular layer reinforcement suture.The da Vinci Xi system was used for surgery, with core instruments including Maryland bipolar forceps (arm 2) and permanent coagulation hook (arm 4). Technical details are provided in Online Resource 1and supplementary materials (Table S1, Text S1).

### 2.4 Statistical Analysis

Data including operative time, intraoperative blood loss, complications (Clavien-Dindo classification), hospital stay, number of harvested lymph nodes, circumferential resection margin status, and R0 resection rate were collected. Functional outcome indicators at 4–8 months postoperatively were evaluated via questionnaires using the Wexner score (fecal incontinence), LARS score (low anterior resection syndrome), IPSS score (International Prostate Symptom Score), and the overall health subscale of EORTC QLQ-C30.Normally distributed continuous variables were expressed as mean ± standard deviation. Non-normally distributed continuous variables were expressed as median (interquartile range). Categorical variables were expressed as frequency (percentage/proportion). All analyses were performed using R 4.5 software.

### 2.5 AI-Assisted Statement

During the writing and revision of this research paper, the authors used artificial intelligence-assisted tools (DeepSeek + Doubao) for language polishing and grammatical optimization to improve the clarity and standardization of academic expression. The use of these tools was limited to text-level assistance. All study design, data collection and analysis, result interpretation, and academic conclusions were independently completed and fully the responsibility of the authors.

## 3. Results

### 3.1 Patient Characteristics and Perioperative Outcomes

A total of 11 patients were included in the analysis (3 males, 8 females), with a mean age of 61.45 ± 8.87 years. All surgeries were completed according to the modular protocol without conversion to open surgery. Perioperative results are shown in Table 1 and Table 2.

**Table 1.**
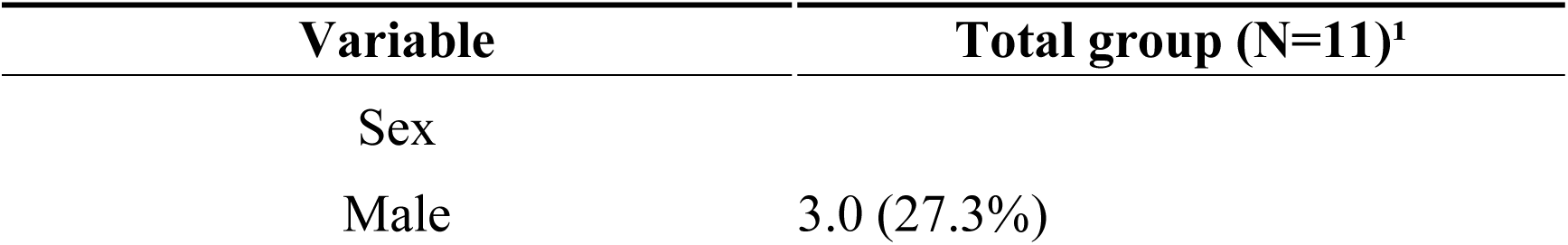
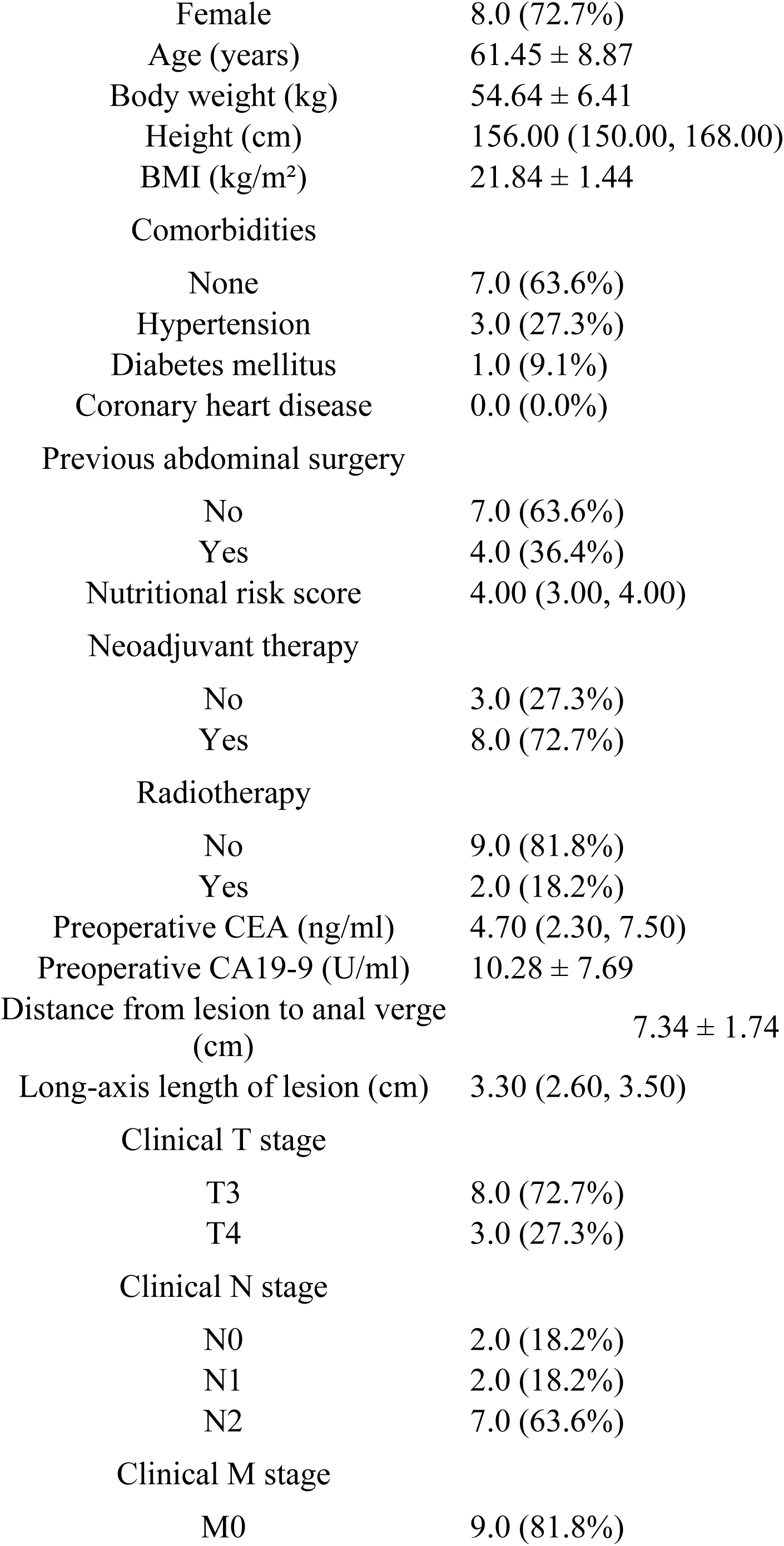
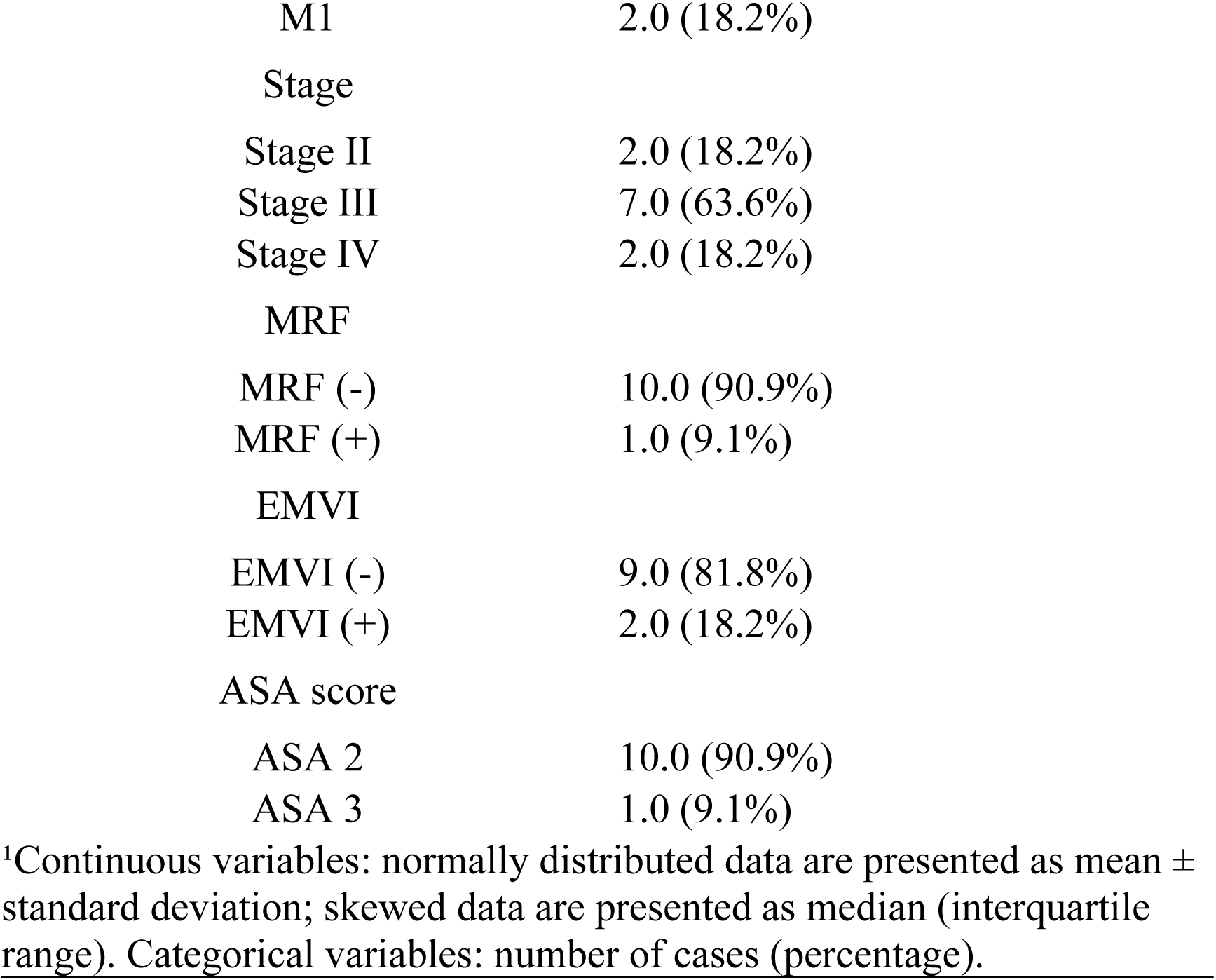
Baseline Characteristics of Patients.

| Variable | Total group (N=11)<sup>1</sup> |
| --- | --- |
| Sex |  |
| Male | 3.0 (27.3%) |
| Female | 8.0 (72.7%) |
| Age (years) | 61.45 ± 8.87 |
| Body weight (kg) | 54.64 ± 6.41 |
| Height (cm) | 156.00 (150.00, 168.00) |
| BMI (kg/m<sup>2</sup>) | 21.84 ± 1.44 |
| Comorbidities |  |
| None | 7.0 (63.6%) |
| Hypertension | 3.0 (27.3%) |
| Diabetes mellitus | 1.0 (9.1%) |
| Coronary heart disease | 0.0 (0.0%) |
| Previous abdominal surgery |  |
| No | 7.0 (63.6%) |
| Yes | 4.0 (36.4%) |
| Nutritional risk score | 4.00 (3.00, 4.00) |
| Neoadjuvant therapy |  |
| No | 3.0 (27.3%) |
| Yes | 8.0 (72.7%) |
| Radiotherapy |  |
| No | 9.0 (81.8%) |
| Yes | 2.0 (18.2%) |
| Preoperative CEA (ng/ml) | 4.70 (2.30, 7.50) |
| Preoperative CA19-9 (U/ml) | 10.28 ± 7.69 |
| Distance from lesion to anal verge (cm) | 7.34 ± 1.74 |
| Long-axis length of lesion (cm) | 3.30 (2.60, 3.50) |
| Clinical T stage |  |
| T3 | 8.0 (72.7%) |
| T4 | 3.0 (27.3%) |
| Clinical N stage |  |
| N0 | 2.0 (18.2%) |
| N1 | 2.0 (18.2%) |
| N2 | 7.0 (63.6%) |
| Clinical M stage |  |
| M0 | 9.0 (81.8%) |

| **Variable** | **Total group (N=11)<sup>1</sup>** |
| --- | --- |
| M1 | 2.0 (18.2%) |
| Stage |  |
| Stage II | 2.0 (18.2%) |
| Stage III | 7.0 (63.6%) |
| Stage IV | 2.0 (18.2%) |
| MRF |  |
| MRF (-) | 10.0 (90.9%) |
| MRF (+) | 1.0 (9.1%) |
| EMVI |  |
| EMVI (-) | 9.0 (81.8%) |
| EMVI (+) | 2.0 (18.2%) |
| ASA score |  |
| ASA 2 | 10.0 (90.9%) |
| ASA 3 | 1.0 (9.1%) |

<sup>1</sup>Continuous variables: normally distributed data are presented as mean ± standard deviation; skewed data are presented as median (interquartile range). Categorical variables: number of cases (percentage).

**Table 2.** Intraoperative and Postoperative Outcomes.

| **Variable** | **Total group (N=11)<sup>1</sup>** |
| --- | --- |
| Operative time (min) | 299.64 ± 54.21 |
| Intraoperative blood loss (ml) | 94.09 ± 50.64 |
| Stoma creation |  |
| No | 11.0 (100.0%) |
| Yes | 0.0 (0.0%) |
| R0 resection |  |
| No | 0.0 (0.0%) |
| Yes | 11.0 (100.0%) |
| Surgical time window |  |
| 8:00–12:00 | 2.0 (18.2%) |
| 12:00–18:00 | 3.0 (27.3%) |
| 18:00–24:00 | 6.0 (54.5%) |
| Differentiation grade |  |
| Well differentiated | 0.0 (0.0%) |
| Moderately differentiated | 9.0 (81.8%) |
| Poorly differentiated | 2.0 (18.2%) |
| Total number of lymph nodes | 17.36 ± 8.44 |
| Number of positive lymph nodes | 0.00 (0.00, 3.00) |
| Number of 253 lymph nodes | 3.09 ± 3.02 |
| Perineural invasion |  |
| Absent | 8.0 (72.7%) |
| Present | 3.0 (27.3%) |
| Vascular invasion |  |
| Absent | 9.0 (81.8%) |
| Present | 2.0 (18.2%) |
| Pathological T stage |  |
| T2 | 2.0 (18.2%) |
| T3 | 9.0 (81.8%) |
| Pathological N stage |  |
| N0 | 6.0 (54.5%) |
| N1 | 4.0 (36.4%) |
| N2 | 1.0 (9.1%) |
| Pathological TRG grade |  |
| 1 | 1.0 (14.3%) |
| 2 | 4.0 (57.1%) |
| 3 | 2.0 (28.6%) |
| Missing | 4 |
| Postoperative hospital stay (days) | 8.2 ± 1.5 |
| Postoperative complications |  |
| No | 8.0 (72.7%) |
| Yes | 3.0 (27.3%) |
| Incomplete intestinal obstruction |  |
| Absent | 8.0 (72.7%) |
| Present | 3.0 (27.3%) |
| Anastomotic leakage |  |
| Absent | 11.0 (100.0%) |

| Variable | Total group (N=11)<sup>1</sup> |
| --- | --- |
| Present | 0.0 (0.0%) |
| Reoperation within 30 days postoperatively |  |
| No | 11.0 (100.0%) |
| Yes | 0.0 (0.0%) |
| Readmission within 30 days postoperatively |  |
| No | 11.0 (100.0%) |
| Yes | 0.0 (0.0%) |

<sup>1</sup>Continuous variables: normally distributed data are presented as mean ± standard deviation; skewed data are presented as median (interquartile range). Categorical variables: number of cases (percentage).

### 3.2 Functional Outcomes

All patients achieved R0 resection. Postoperative patient-reported outcome (PRO) scores are shown in Table 3.

**Table 3.** Quality of Life and Functional Scores.

| Variable | Total group (N=11)<sup>1</sup> |
| --- | --- |
| Wexner fecal incontinence score | 4.18 ± 2.89 |
| Wexner constipation score | 3.27 ± 2.45 |
| LARS score | 25.36 ± 7.97 |
| Urinary function | 3.36 ± 3.85 |
| Physical function | 98.18 ± 4.31 |
| Role function | 96.97 ± 10.05 |
| Emotional function | 100.00 ± 0.00 |
| Cognitive function | 98.48 ± 5.03 |
| Social function | 96.97 ± 6.74 |
| Global health status | 87.88 ± 7.79 |
| Fatigue | 7.07 ± 16.68 |
| Nausea and vomiting | 0.00 ± 0.00 |
| Pain | 9.09 ± 8.70 |
| Dyspnea | 3.03 ± 10.05 |
| Insomnia | 12.12 ± 16.82 |
| Appetite loss | 0.00 ± 0.00 |
| Constipation | 15.15 ± 22.92 |
| Diarrhea | 34.85 ± 28.34 |
| Financial difficulties | 21.21 ± 16.82 |

<sup>1</sup>All variables are continuous variables, expressed as mean ± standard deviation

## 4. Discussion

The integration of robotic surgical systems and natural orifice specimen extraction (NOSES) is advancing rectal cancer surgery toward greater precision and minimal invasiveness. However, this technique is operationally complex and lacks a standardized framework, which restricts its clinical promotion and standardized teaching[23]. This study is the first to systematically propose and preliminarily validate a set of structured modular robotic NOSES-II surgical workflows for mid-rectal cancer. Its core innovation lies in the construction of a technical system consisting of six logical modules, which integrates the principles of “plane priority and nerve preservation” with the NOSES goal of “no incision”, and translates them into concrete steps that can be standardized for implementation and teaching, providing a systematic solution to overcome the challenges of steep learning curve and inconsistent operation of this procedure.

This structured workflow has demonstrated good safety and feasibility in preliminary application, with all surgeries completed smoothly according to the modules and no conversion to open surgery. The mean operative time (299.6 ± 54.2 minutes) was longer than that reported in the study by LIU et al.[10], which may be related to the initial stage of the surgeon’s learning curve and the higher operational difficulty of lower tumor location. The mean intraoperative blood loss was similar, reflecting the technical advantages of the robotic system in achieving precise dissection and stable hemostasis within the narrow pelvis, which guarantees the precise implementation of modular operations. More importantly, while pursuing minimal invasiveness, this protocol strictly follows the principle of oncological radicality, achieving a 100% R0 resection rate and adequate lymph node dissection, confirming the synergy between structured operation and high-quality total mesorectal excision (TME).

The hallmark feature of this surgery is the completion of specimen extraction through the natural orifice, which fundamentally avoids abdominal auxiliary incisions and stomas. This not only preserves the patient’s somatic integrity but also completely eliminates stoma-related complications and long-term physical and psychological burdens[24]. Although preventive stoma is not routine for mid-rectal cancer, clinical decision-making is often influenced by the uncertainty of intraoperative assessment[25,26]. Through precise modular operations (such as tension-free anastomosis and ICG blood supply assessment), this study transforms “stoma avoidance” from a limited compromise option into a determinable surgical goal that can be actively achieved, providing a new technical path for improving patients’ postoperative quality of life.

Early postoperative patient-reported outcomes (PROs) showed that low Wexner scores and IPSS scores indicated good recovery of defecatory and urinary function in patients. This benefits from the proactive nerve preservation strategy embedded in the protocol, that is, guiding plane expansion with clear fascial landmarks, and identifying and preserving autonomic nerve plexuses at key steps, transforming nerve preservation from empirical operation into a module that can be standardized for implementation. In addition, the efficient combination of Maryland bipolar forceps and permanent coagulation forceps is suitable for modular fine dissection, and its cost-effectiveness is also conducive to the promotion of this technique.

This study has certain limitations: it is a preliminary summary of a single-center, small sample, and the conclusions need to be verified by larger-scale studies; the lack of a prospective randomized controlled design makes it difficult to quantitatively evaluate the advantages of this protocol over other procedures in terms of long-term oncological outcomes and functional benefits. Future prospective comparative studies are needed to focus on evaluating the value of this structured procedure in shortening the learning curve, improving long-term quality of life, and health economic benefits. The accompanying high-definition surgical videos and technical documents can provide important support for technical standardization, peer review, and teaching promotion.

Currently, surgery is moving toward digitalization and intelligence[27–29]. The structured procedure and video database established in this study lay the foundation for integration with artificial intelligence (AI) and computer vision technologies. The use of AI for automatic analysis of surgical videos is expected to realize real-time objective assessment of the quality of each operation module, promoting the transformation of surgical training and quality management from experience-driven to data-driven models[30,31]. This is not only a technical innovation but also provides a structured starting point for the development of the next generation of intelligent and standardized rectal cancer surgery.

## 5. Conclusion

Preliminary studies confirm that the structured modular robotic NOSES-II surgical procedure is safe and feasible for the treatment of mid-rectal cancer. By providing a clear technical framework and systematic integration of nerve preservation, this procedure establishes a standardized operational path for this complex minimally invasive procedure, and provides a practical solution for standardized clinical implementation and teaching promotion.

## Supporting information

Supplementary video Online Resource 1 Figure S1 Table S1 andText S1

## Data Availability

All data produced in the present study are available upon reasonable request to the authors

## Supplementary Materials Description

- **Online Resource 1:** Full procedure demonstration video of the six-step structured modular robotic NOSES-II surgery. Duration: [00:09:16].
- **Figure S1:** Schematic diagram of surgical setup and core instruments.
- **Table S1:** Technical summary table of the six-module structured surgical procedure.
- **Text S1:** Step-by-step detailed technical analysis of the structured surgical procedure.

