## Supplementary video Online Resource 1 Figure S1 Table S1 andText S1 for "Modular Robotic NOSES-II for Mid-Rectal Cancer: A Preliminary Feasibility Study of a Structured Surgical Procedure"

**Corresponding Article**

**Online Resource 1**

**Title:** Full Procedure Demonstration of Six-Step Modular Robotic NOSES-Ⅱ Surgery

**Video Caption:** This video fully demonstrates the modular robotic NOSES-Ⅱ surgery for mid-rectal cancer, strictly following the six predefined modules described in the main manuscript. Key steps highlighted include:

- **Module 1: Medial approach and establishment of the safe dorsal plane:** Enter and expand Toldt's space and the retrorectal space, with the anterior fascia of the hypogastric nerve as the dorsal landmark.
- **Module 2: Refined IMA management and nerve preservation:** Perform D3 lymph node dissection at the root of the IMA, and ligate the IMA at a low position distal to the origin of the left colic artery (LCA).
- **Module 3: Mobilization of the left colon and splenic flexure:** Complete mobilization of the left colon via a combined medial and lateral approach.
- **Module 4: Fluorescence-guided mesenteric tailoring:** Perform skeletonized tailoring of the sigmoid mesocolon guided by indocyanine green fluorescence imaging to assess blood supply.
- **Module 5: Nerve-preserving circumferential rectal dissection:** Perform total mesorectal excision within the "holy plane" in the order of "posterior → lateral → anterior".
- **Module 6: Transanal specimen extraction and reconstruction:** Complete transanal specimen extraction and intracorporeal anastomosis using the NOSES-Ⅱ B technique.

The video is accompanied by voiceover narration and subtitles for key steps, highlighting important anatomical landmarks, technical essentials, and principles of neurovascular preservation.

**Duration:** [09 min 16 sec]

**Supplementary Figure S1**

**Title:** Surgical Setup and Core Instruments

**Figure Legend:**

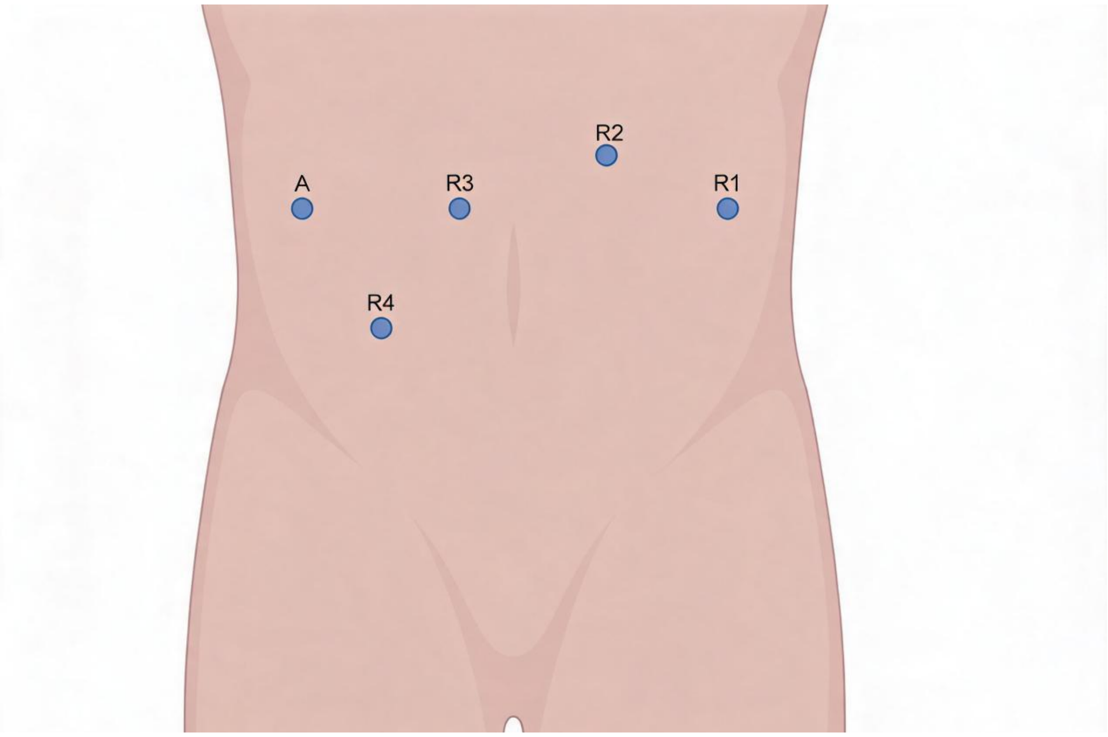


(A) Abdominal trocar placement diagram. The diagram shows the positions of robotic ports: R1 (Cadiere forceps), R2 (Maryland bipolar forceps), R3 (camera), R4 (permanent coagulation hook), and the assistant port (A).

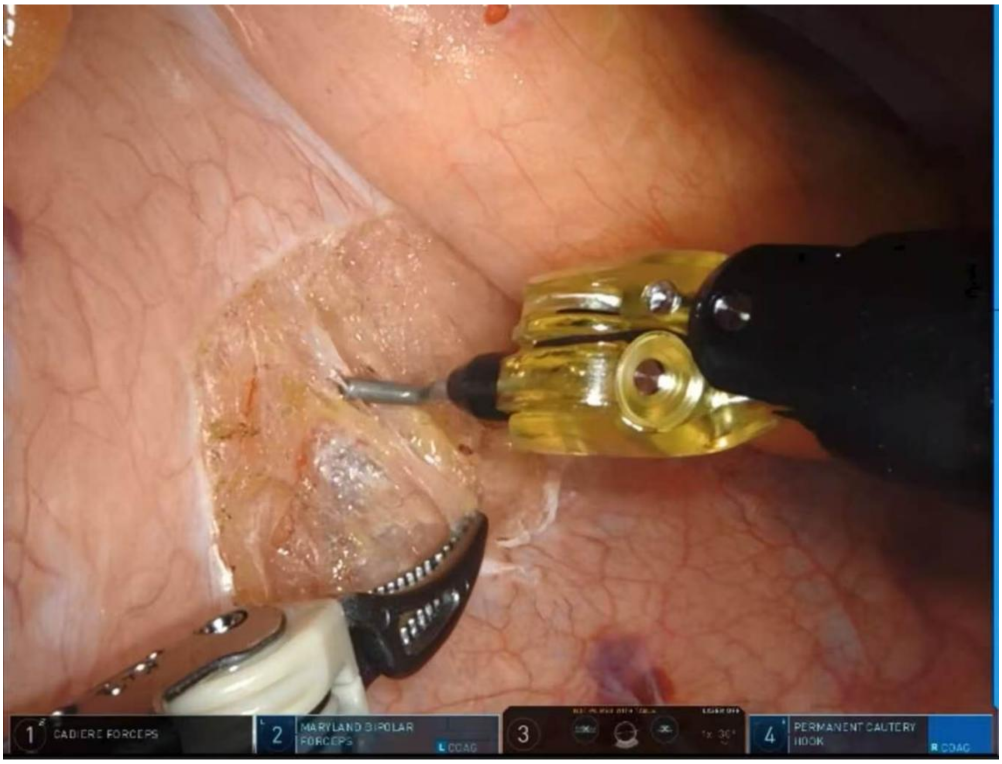


(B) Close-up view of core robotic instruments: Maryland bipolar forceps and permanent coagulation hook.

**Supplementary Table S1**

**Title:** Technical Summary Table of the Six-Module Structured Surgical Procedure

| Module | Core Objective | Key Anatomical Planes/Structures | Instrument Synergy and Key Maneuvers | Key Points of Nerve Preservation |
|---|---|---|---|---|
| 1. Medial approach | Establish a safe dorsal plane encompassing Toldt's space and the retrorectal space. | Anterior fascia of the hypogastric nerve, superior hypogastric plexus | R1 lifts the mesentery, R2 assists in exposure, and R4 performs sharp dissection. | Expand the plane anterior to the anterior fascia of the hypogastric nerve to preserve the superior hypogastric plexus. |
| 2. IMA management | Perform D3 lymph node dissection, and achieve low ligation of the IMA with LCA preservation. | IMA vascular sheath, left colic artery (LCA), inferior mesenteric plexus | R2 retracts the vascular sheath, and R4 performs precise extra-sheath dissection. | Treat perforating branches 2–3 mm away from the main vascular trunk to preserve the inferior mesenteric plexus and LCA branches. |
| 3. Left colon mobilization | Completely mobilize the left colon to the splenic flexure. | Toldt's fascia, left ureter, gonadal vessels | R2 provides multi-directional traction, and R4 coagulates tiny vessels. | Maintain the integrity of the prerenal fascia, and identify and preserve the ureter and gonadal vessels under direct vision. |
| 4. Mesenteric tailoring | Precisely divide the marginal vascular arcade to ensure blood supply to the anastomosis. | Sigmoid vascular arcade, marginal artery | ICG fluorescence confirms perfusion, and R4 precisely divides vascular branches. | Ensure blood supply to the reserved intestinal segment to avoid ischemic nerve injury. |
| 5. Rectal dissection | Perform total mesorectal excision and preserve pelvic autonomic nerves. | Rectal proper fascia ("holy plane"), Denonvilliers' fascia, pelvic plexus, neurovascular bundle (NVB) | The "triangular traction" technique is adopted, with R2 and R4 operating within the fascial interface. | Posteriorly along the "holy plane"; laterally preserve the pelvic plexus; anteriorly preserve the NVB posterior to |

| Module | Core Objective | Key Anatomical Planes/Structures | Instrument Synergy and Key Maneuvers | Key Points of Nerve Preservation |
|---|---|---|---|---|
| | | | | Denonvilliers' fascia. |
| 6. Specimen extraction and reconstruction | Extract the specimen transanally and complete tension-free anastomosis. | Anal canal, anastomosis | The NOSES- II B technique (intracorporeal division) is adopted, and the stapler is inserted transanally. | Gentle manipulation to avoid sphincter injury and ensure good blood supply to the anastomosis. |

**Supplementary Text S1**
**Title:** Step-by-Step Technical Analysis of the Structured Surgical Procedure
This text provides a detailed technical analysis of the six surgical modules, serving as a textual supplement to Supplementary Video 1 and Supplementary Table S1. The content elaborates on the refined operational steps, key anatomical identification points, and operational principles of each module.

**Module 1: Medial Approach and Anatomical Plane Establishment**
This module aims to establish a clear anatomical plane throughout the abdominopelvic cavity under bloodless conditions via the medial approach, and systematically preserve the pelvic autonomic nerve structures. The anterior fascia of the hypogastric nerve serves as the key dorsal anatomical landmark and protective interface.

**Surgical steps:** The posterior peritoneum is incised approximately 1 cm below the projection of the inferior mesenteric artery to enter the loose layer at the junction of Toldt's space and the retrorectal space. The assistant provides counter-traction to maintain tension. The surgeon uses Maryland bipolar forceps (arm R2) for exposure and a permanent coagulation hook (arm R4) for precise sharp dissection. Dissection is performed on the superficial side of the anterior fascia of the hypogastric nerve, along the "holy plane" between it and the rectal proper fascia. First, expand the retrorectal space caudally, then expand Toldt's space cranially and to the left. Maintain the integrity of the rectal proper fascia throughout the procedure, ensuring all dissection is performed on the superficial side of the anterior fascia of the hypogastric nerve. A safe and continuous abdominopelvic anatomical plane is established to protect the underlying superior hypogastric plexus, bilateral hypogastric nerves, and pelvic autonomic nerves, laying an anatomical foundation for subsequent procedures.

**Module 2: Management of the Inferior Mesenteric Artery Region and Lymph Node Dissection**
This module aims to precisely complete D3 lymph node dissection in the IMA region, perform low ligation of the IMA with LCA preservation, and preserve the inferior mesenteric plexus throughout the procedure.

**Surgical steps:**

**D3 lymph node dissection (group 253):** En bloc resection of lymphoadipose tissue outside the vascular sheath is performed along the root of the IMA. The dissection is advanced step by step in the order of "medial boundary → cranial boundary → caudal boundary → lateral boundary → dorsal boundary". Gently lift the tissue with Maryland bipolar forceps, and use the coagulation hook to precisely treat perforating vessels 2–3 mm away from the main vascular trunk. When dissecting the dorsal boundary, pull the proximal IMA ventrally for adequate exposure.After confirming the course of the LCA, determine the ligation plane distal to its origin. Skeletonize the IMA vessel, close the proximal end with absorbable vascular clips and then divide it. The left colic artery is preserved, and the IMA is ligated at a low position.Pay attention to preserving the inferior mesenteric plexus throughout the procedure. The high-magnification field of view and stable manipulation of the robotic system provide ideal support for LCA identification, refined IMA ligation, and nerve preservation.

This module achieves radical D3 dissection, ensures intestinal blood supply by preserving the LCA, and systematically protects the inferior mesenteric plexus to maintain postoperative function.

**Module 3: Mobilization of the Left Colon and Splenic Flexure**

Via a combined approach of the medial Toldt's space and the lateral paracolic sulcus, this module achieves adequate mobilization of the left colon and splenic flexure, provides conditions for tension-free anastomosis, and protects important left retroperitoneal structures.

**Surgical steps:** Lift the peritoneum of the descending mesocolon, expand Toldt's space cranially and laterally along the superficial side of the prerenal fascia, and coagulate tiny vessels. Retract the colon medially, and sharply divide the lateral peritoneum cranially along the "yellow-white junction line" to the splenocolic ligament. Individualize the release of the splenocolic ligament according to the length required for anastomosis, and divide part of the gastrocolic ligament if necessary to achieve sufficient release. All medial dissection is performed on the superficial side of the prerenal fascia, which acts as a natural barrier to protect the underlying left ureter and gonadal vessels. Pay attention to preserving the colonic marginal vascular arcade during the procedure.

Complete release of the left colon is achieved, ensuring that the proximal colon can be pulled down to the pelvic cavity without tension, and the left ureter, gonadal vessels, and marginal vascular arcade are fully preserved.

**Module 4: Colonic Mesenteric Tailoring and Blood Supply Assessment**

This module completes precise tailoring and blood supply optimization of the proximal colonic mesentery, laying the foundation for tension-free anastomosis with good blood perfusion.

**Surgical steps:** Guided by the descending branch of the left colic artery, precisely tailor the sigmoid mesocolon toward the proximal pretransection plane at approximately 1 cm from the marginal vascular arcade. Indocyanine green fluorescence imaging is routinely or selectively applied to clearly visualize the course of the marginal vascular arcade and evaluate the perfusion of the reserved intestinal segment. After confirming blood supply, selectively divide the vascular branches corresponding to the intestinal segment to be resected, and finally completely divide the mesentery and marginal vascular arcade at the proximal colon pretransection plane. Always maintain a safe distance from the marginal vascular arcade, and adequate blood supply to the reserved intestine must be confirmed before final division.

Mesenteric tailoring conforming to anatomical layers is achieved, and reliable blood supply to the reserved intestinal segment is ensured through intraoperative assessment, making full preparation for anastomosis.

**Module 5: Circumferential Rectal Dissection and Autonomic Nerve Preservation**

Under the principle of total mesorectal excision, this module completes anatomical circumferential dissection of the rectum and systematically implements pelvic autonomic nerve preservation.

**Surgical steps:** Traction the rectum ventrally, and sharply dissect along the interface between the rectal proper fascia and the anterior fascia of the hypogastric nerve to the predetermined distal resection margin. Use the "triangular traction" technique to expose the pelvic sidewall, precisely dissect and divide the lateral rectal ligaments at the plane medial to the pelvic nerve plexus, and preserve the pelvic nerve plexus and inferior hypogastric nerves. Dissect along the plane posterior to Denonvilliers' fascia; in males, preserve the prostatic capsule and neurovascular bundle; in females, pay attention to preserving the posterior vaginal wall.

Adhere to the "plane priority" principle, with all dissection performed within definite anatomical interfaces; implement "nerve-guided" active preservation using nerve structures as key landmarks; maintain the integrity of the rectal proper fascia to ensure a negative circumferential resection margin.

Radical resection conforming to TME principles is achieved, pelvic autonomic nerves are maximally preserved to retain urogenital function, and the resection range is individualized according to tumor location.

**Module 6: Specimen Extraction and Digestive Tract Reconstruction**

Using the NOSES- II B technique, this module completes specimen extraction and digestive tract reconstruction via the natural orifice, achieving zero abdominal wall incisions.

**Surgical steps:** Skeletonize the intestinal wall ≥5 cm distal to the tumor and transect the rectum with a linear cutter stapler. Divide the proximal colon and perform purse-string suture on the cut end. After anal dilation, make an incision at the staple line on the anterior wall of the rectal stump. Pull the distal end of the sterile specimen protection bag inserted through the main operating port out through the anus, and deliver the circular stapler anvil into the abdominal cavity through the bag. Place the specimen into the bag and extract it completely through the anus. Implant and fix the anvil into the cut end of the proximal colon. Insert the main body of the circular stapler transanally to complete colo-rectal end-to-end anastomosis, and perform circumferential seromuscular reinforcement suture. Evaluate anastomotic blood supply via fluorescence imaging. Place a drainage tube at the pelvic floor and retain an anal tube for decompression transanally.

Strictly abide by the aseptic and tumor-free principles when using the protection bag; routinely evaluate blood supply after anastomosis; ensure the anastomosis is tension-free.
Minimally invasive transnatural orifice specimen extraction is achieved, functional digestive tract reconstruction with good blood supply and no tension is completed, and anastomotic healing is supported through drainage and decompression measures.
